# Proximity of Wildfires to Inpatient Healthcare Facilities in California, 2001-2023

**DOI:** 10.1101/2025.02.15.25322338

**Authors:** Caleb Dresser, Neil Singh Bedi, Andrew Schroeder, Eric Sergienko, Satchit Balsari

**Affiliations:** Department of Emergency Medicine, Beth Israel Deaconess Medical Center, Boston, MA; Harvard Center for Climate, Health, and the Global Environment, Department of Environmental Health, Harvard TH Chan School of Public Health, Boston, MA; Harvard T.H. Chan School of Public Health, Boston, MA; Boston University Chobanian and Avedisian School of Medicine, Boston, MA; Direct Relief, Santa Barbara, USA; Former Health Officer, Mariposa County Health and Human Service Agency; Department of Global Health and Population, Harvard T.H. Chan School of Public Health

**Keywords:** wildfire, hospital, spatial, disaster, preparedness, climate

## Abstract

**Objective:** Assess changes in the proximity of wildfires to inpatient healthcare facilities in California during the period 2001 to 2023.

**Methods:** Retrospective spatial analysis. Distances between each inpatient facility and the nearest wildfire perimeter were computed on an annual basis using data from the California Department of Health Care Access and Information and CAL-FIRE’s Fire and Resource Assessment Program. Temporal changes were analyzed via Kruskal-Wallis and linear models.

**Results:** The distance from inpatient healthcare facilities in California to nearby wildfires is decreasing by an average of 628 feet per year, while close approaches are increasing; during 2017-2023, there was 53% increase in the number of inpatient beds within five miles of a wildfire as compared with 2001-2008.

**Conclusion:** Wildfires are occurring closer to inpatient healthcare facilities in California. An increasing proportion of California’s inpatient bed capacity is exposed to nearby wildfires. Policies to reduce risk posed by wildfires, prepare for evacuations, preserve access to healthcare, and ensure safe location of new facilities are urgently needed to ensure the safety of patients and the wellbeing of populations that depend on inpatient healthcare services.

## INTRODUCTION

Wildfires in California are becoming larger and more destructive.^1^ Worsening wildfire risks related to factors including changing development and land use patterns, human activity and related ignition sources, and the impacts of global climate change.^2^ Of the 20 largest fires in state history, 13 took place in 2015 or later.^3^ Fifteen of the 20 most destructive wildfires took place in the same time period.^3^

Wildfires and their smoke affect human health through both direct and indirect pathways. In addition to causing respiratory illnesses, impacts of wildfire smoke on patients with cardiovascular or respiratory disease and other conditions are also well documented.^4-7^ Evacuations, property damage, and disruptions in critical infrastructure, social functioning and economic activity have also been shown to result in negative impacts on health.^8-10^ Health systems can suffer both direct damage to their structures and indirect effects related to disruption of critical infrastructure, transportation networks, supply chains, and population displacement, including staff evacuations.^11,12^

A large proportion of the inpatient bed capacity in California is located near fire threat zones described as “high”, “very high”, or “extreme” by the California Department of Forestry and Fire Protection (CAL-FIRE), with half of California’s inpatient bed capacity located less than 0.87 miles from a high fire threat zone in 2022.^13^ However, empirical information on how this risk translates into actual exposure of healthcare facilities to wildfires, information on temporal trends in their exposure, and information on spatial distribution of such exposures are lacking. This information is important for assessing vulnerability, identifying adaptation opportunities, and prioritizing finite resources toward settings where they will be maximally beneficial.

In this analysis, we assess wildfire proximity to critical healthcare infrastructure in California, specifically inpatient healthcare facilities. This retrospective spatial analysis provides information on the changing proximity of wildfires to inpatient healthcare facilities in California during the period 2001-2023.

## DATA & METHODS

This study consisted of a retrospective geospatial analysis of distances from inpatient healthcare facilities in California to the nearest wildfire in each year using publicly available data on healthcare facilities and historical wildfire perimeters (Supplement A).

Healthcare Facility Data: Inpatient healthcare facility data (2001-2023) was sourced from the California Department of Health Care Access and Information.^14^ Inpatient facilities include general acute care hospitals, psychiatric hospitals, chemical dependency recovery hospitals, skilled nursing facilities, intermediate care facilities, congregate living health facilities, and hospice facilities, comprising all facilities where patients require overnight care and could require evacuation in the event of a disaster. Facilities that had suspended licenses, were closed, or were otherwise non-operational were excluded from analysis in the applicable years.

Wildfire Perimeter Data: Data on wildfire perimeters was retrieved from the CAL-FIRE Fire and Resource Assessment Program (FRAP).^15^ Wildfire perimeter data is based on information from ground teams, aerial surveillance, and satellite imagery. Although CAL-FIRE FRAP hosts “the most complete digital record of fire perimeters in California," some fires may be missing, may have been excluded due to minimum cutoffs, may be insufficiently or inaccurately documented, or have not yet been added to the database.^15^

Distance Calculations: The distance from each facility that was open in a given year to the nearest point or vector in any wildfire perimeter from that year was computed using the sf package in R; this process was repeated separately for each year in which the facility was open.^16^ A single numerical value representing the distance from the facility to the nearest wildfire perimeter (‘wildfire-facility distance’; Supplement B) was computed for each facility for each year. In cases where facilities were within the fire perimeter, distances were coded as zero.

Analysis: A Kolmogorov-Smirnov test was performed to assess the normality of the distribution of the distances in each year. The dataset was subdivided into three time periods (tertiles) by year, specifically 2001-2008, 2009-2016, and 2017-2023. Differences in distances from facilities to wildfires were assessed by Kruskal Wallis tests, due to the non-normal distribution of wildfire distances within each year. A Dunn test was conducted, and differences of medians were computed between each pairing of the three tertiles. Cumulative inpatient bed capacity percentage plots relative to distance to nearest wildfire were constructed for the three time periods. A simple linear regression was also performed, taking wildfire-facility distances as a response variable and year as a predictor variable. The purpose of this regression was to confirm the overall trend in distances from facilities to wildfires over time; causal inference was not attempted.

A sub-analysis of the late study period (2017-2023) was performed to assess whether more recently opened facilities were disproportionately contributing to the lower wildfire-facility distances in the late period; a Wilcoxon rank sum test was conducted to compare wildfire-facility distances of facilities that were first licensed and opened in or after 2017 against those licensed prior to 2017.

All analyses were completed using R (Version 4.3.1) in RStudio (Version 2024.04.2+764) and Microsoft Excel (Version 16.86). This study utilized publicly available data from the State of California, did not involve human subjects research, and did not require IRB review. The analysis code is available in a public GitHub repository (https://github.com/nsbedi/wildfire-distances-inpt-hcfs).

## RESULTS

The annual number of inpatient healthcare facilities ranged from 1,588 in 2002 to 1,867 in 2020, with a modest upward trend in the total number of facilities during the study timeframe (Table 1). Inpatient healthcare facilities included general and acute care hospitals and inpatient psychiatric hospitals (“hospitals”), which accounted for 492 to 518 facilities per year, as well as long term care facilities (“LTCFs”), which accounted for 1,070 to 1,367 facilities per year. Growth in the total number of facilities over time was predominantly due to the increasing number of LTCFs (Table 1).

**Table 1:** Inpatient Healthcare Facilities and Wildfires in California, 2001-2023.

| Year | Hospitals | LTCFs | Total Facilities | Number of Fires <sup>a</sup> | Acres Burned <sup>a,b</sup> | Median Wildfire-Facility Distance (miles) |
| --- | --- | --- | --- | --- | --- | --- |
| 2001 | 512 | 1202 | 1714 | 206 | 246,925 | 17.24 |
| 2002 | 518 | 1070 | 1588 | 243 | 963,899 | 12.27 |
| 2003 | 509 | 1135 | 1644 | 339 | 968,697 | 14.12 |
| 2004 | 501 | 1194 | 1695 | 277 | 274,639 | 14.94 |
| 2005 | 500 | 1180 | 1680 | 304 | 231,112 | 14.19 |
| 2006 | 502 | 1188 | 1690 | 314 | 748,170 | 14.61 |
| 2007 | 503 | 1187 | 1690 | 348 | 1,031,803 | 9.5 |
| 2008 | 502 | 1189 | 1691 | 439 | 1,382,710 | 10.64 |
| 2009 | 499 | 1192 | 1691 | 254 | 435,825 | 12.14 |
| 2010 | 501 | 1191 | 1692 | 207 | 101,056 | 14.31 |
| 2011 | 502 | 1193 | 1695 | 316 | 202,423 | 14.44 |
| 2012 | 500 | 1199 | 1699 | 352 | 848,692 | 12.78 |
| 2013 | 498 | 1200 | 1698 | 297 | 569,422 | 14.42 |
| 2014 | 500 | 1215 | 1715 | 236 | 554,656 | 16.42 |
| 2015 | 497 | 1248 | 1745 | 317 | 789,222 | 10.6 |
| 2016 | 497 | 1266 | 1763 | 361 | 546,250 | 11.73 |
| 2017 | 492 | 1324 | 1816 | 608 | 1,424,583 | 7.69 |
| 2018 | 497 | 1338 | 1835 | 416 | 1,581,917 | 10.5 |
| 2019 | 493 | 1353 | 1846 | 319 | 280,728 | 9.75 |
| 2020 | 493 | 1374 | 1867 | 505 | 4,182,335 | 8.83 |
| 2021 | 496 | 1365 | 1861 | 388 | 2,512,611 | 9.85 |
| 2022 | 498 | 1367 | 1865 | 306 | 322,528 | 10.06 |
| 2023 | 500 | 1347 | 1847 | 284 | 342,033 | 18.21 |

<sup>a</sup> Data sourced from CAL-FIRE FRAP. <sup>b</sup> Rounded to the nearest whole acre.

In the period 2001-2023, CAL-FIRE FRAP recorded 22,261 fires, with annual burned acreage ranging from 101,056 to 4,182,335 acres (Table 1). There was an increase in the number of acres burned per year during the study timeframe; four of the six years in which more than 1,000,000 acres burned occurred in the second half of the analysis period.

A total of 40,027 wildfire-facility distances were computed over the 23-year study timeframe. The result was a single dataset of operational inpatient healthcare facilities in California in each year (2001-2023) and the distance from each facility to the nearest wildfire perimeter recorded by CAL-FIRE that year. This dataset has been made publicly available in a data repository (https://doi.org/10.7910/DVN/MODVUK).

### Distribution of Wildfire-Facility Distances

Distances from inpatient healthcare facilities to the nearest wildfire perimeter ranged from 0 to 167.7 miles; annual median wildfire-facility distances ranged from 7.69 to 18.21 miles (Table 1). Wildfire-facility distances in each year were asymmetrically distributed with a rightward skew, reflecting the subset of facilities that did not experience any nearby fires in that year (Figure 1). The data was non-normally distributed in all years, as assessed by the Kolmogorov-Smirnov test (P < 0.001 for all years); the shape of the distributions was relatively consistent across years (Supplement C).

**Figure 1:**
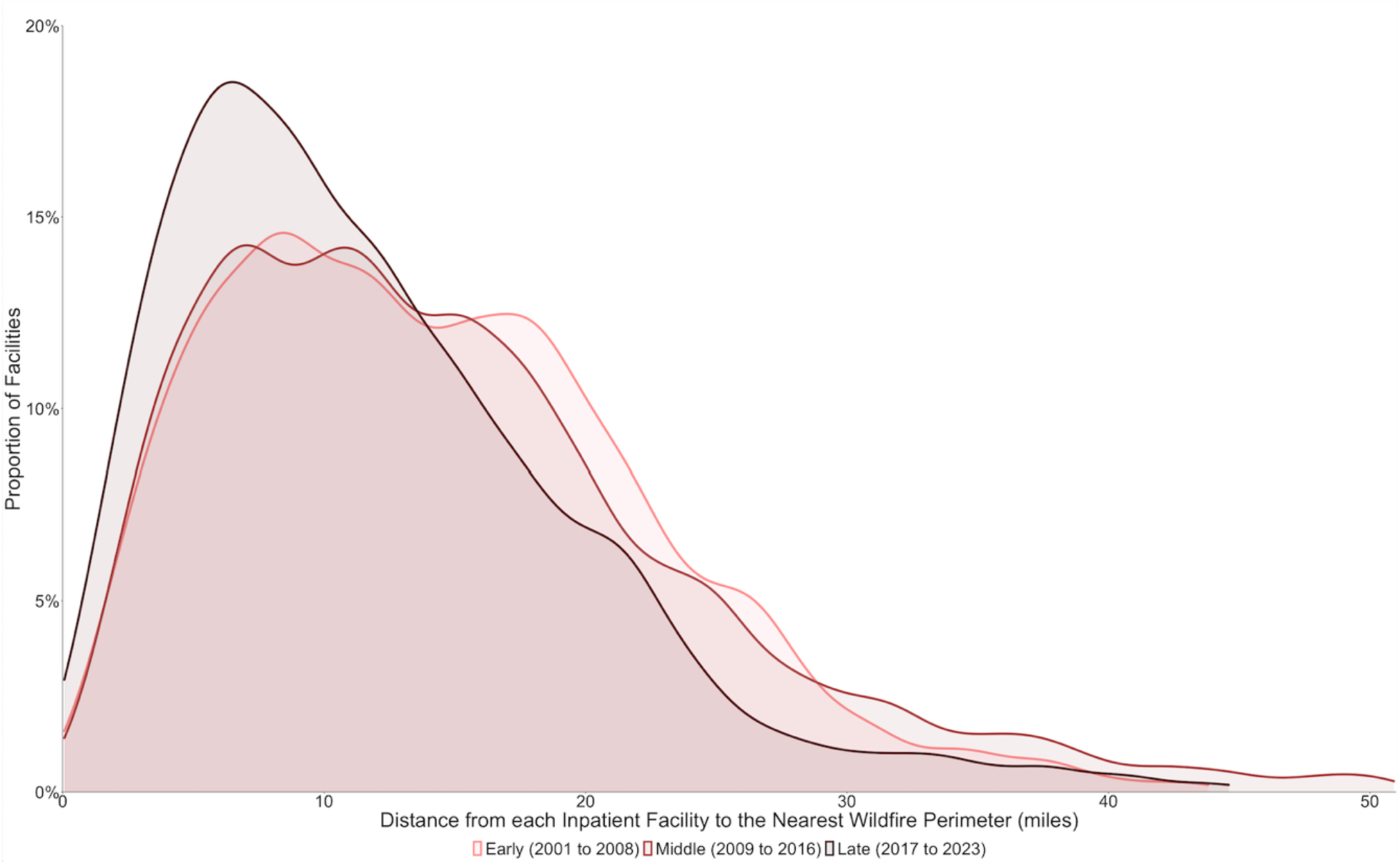
Wildfire-Facility Distance Distributions in the Early, Middle, and Late Time Periods. Distribution of wildfire-facility distances for the Early (2001-2008), Middle (2009-2016), and Late (2017-2023) time periods. Wildfires were closer to healthcare facilities in the late period as compared with the early and middle time periods.

### Temporal Changes in Wildfire Facility Distances

To assess change in wildfire-facility distances over time, the dataset was subdivided into temporal tertiles: ‘early’ (2001-2008), ‘middle’ (2009-2016), and ‘late’ (2017-2023). Kruskal-Wallis confirmed a statistically significant difference in wildfire-facility approach distances between time periods (H=952.8, df=2, P<0.0001), and a Dunn test confirmed the significance of differences between individual subsets (P< 0.0001 for late-early and late–middle). There were lower median wildfire-facility distances in the late period (10.32 miles, IQR 10.12) as compared with the early (13.33 miles, IQR 11.55) and middle (13.27 miles, IQR 12.20) time periods. A higher proportion of inpatient healthcare facilities experienced nearby wildfires in the late period as compared with the early and middle time periods (Figure 2).

**Figure 2:**
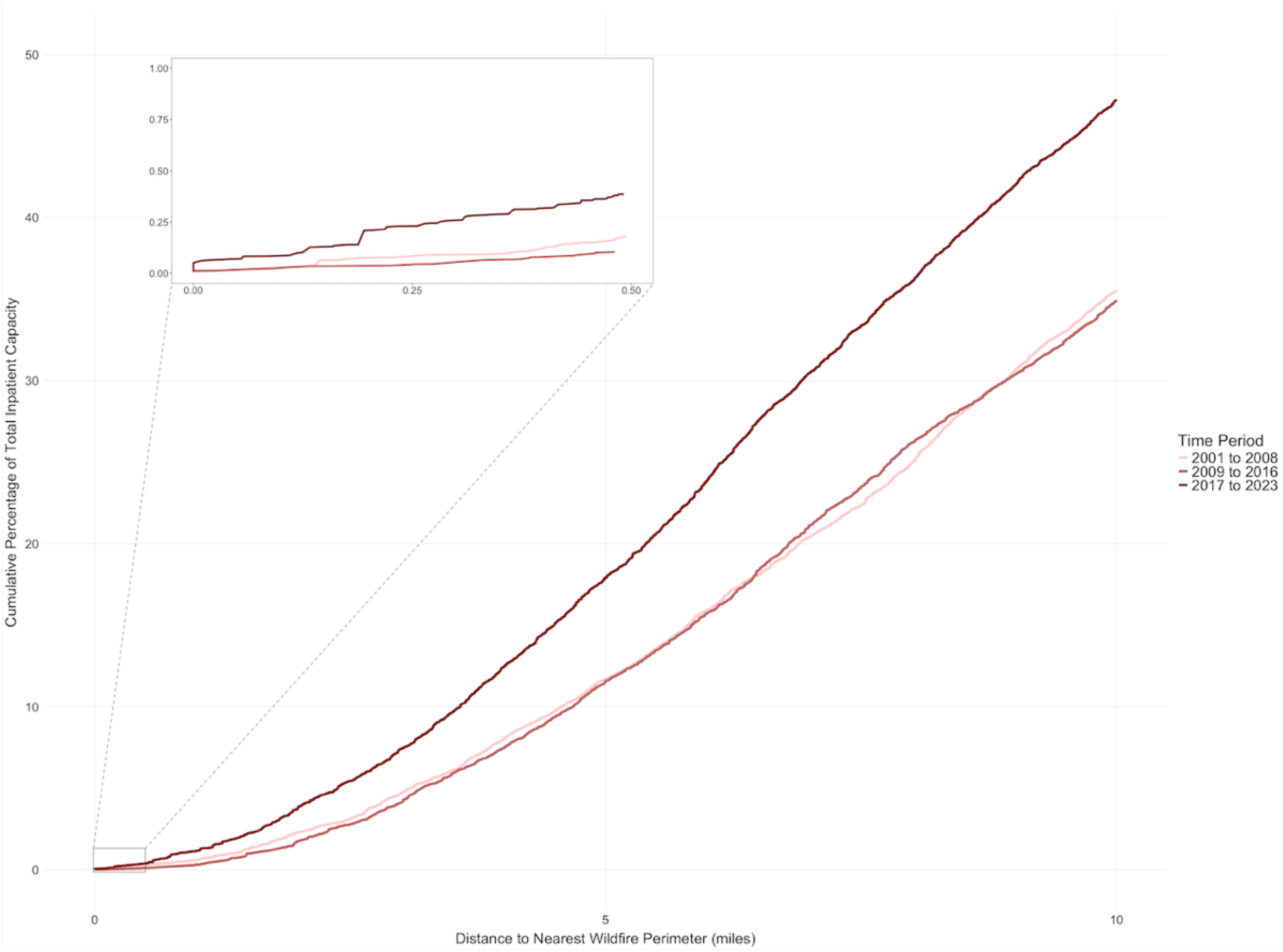
Cumulative Percentage of Total Inpatient Bed Capacity Exposed to Nearby Wildfires in the Early, Middle, and Late Time Periods. Total inpatient bed capacity was aggregated for the Early (2001-2008), Middle (2009-2016), and Late (2017-2023) time periods based on proximity to the nearest wildfire perimeter and is presented via cumulative percentage plots. The proportion of inpatient healthcare capacity within five miles of a wildfire approach was 53% higher in the most recent (Late) period as compared to previous time periods.

A univariate linear model was constructed to confirm the relationship between wildfire-facility distances and time. This model incorporated distances as a response variable and year as a predictor variable; no attempt was made to adjust for covarying factors, as the goal was to describe the trend in observed data rather than make causal inferences. In this model, the passage of time was associated with a statistically significant reduction in wildfire-facility approach distances of approximately 0.128 miles (678 feet; 206 meters) per year (P < 0.0001). The overall explanatory value of the model for this system was poor (R^2^ = 0.008), as was expected given the univariate approach, the stochasticity of wildfire location each year, and the decision to use a descriptive rather than causal or predictive modeling approach.

Total inpatient bed capacity for each of the three time periods was aggregated by proximity to the nearest wildfire perimeter and is presented in cumulative percentage plots (Figure 3). The proportion of inpatient bed capacity that experienced the occurrence of a wildfire within five miles was 53% higher in the late period (2017-2023) than in the early period (2001-2008). Change in exposure in the late period relative to the middle period showed a similar increase (Figure 3).

**Figure 3:**
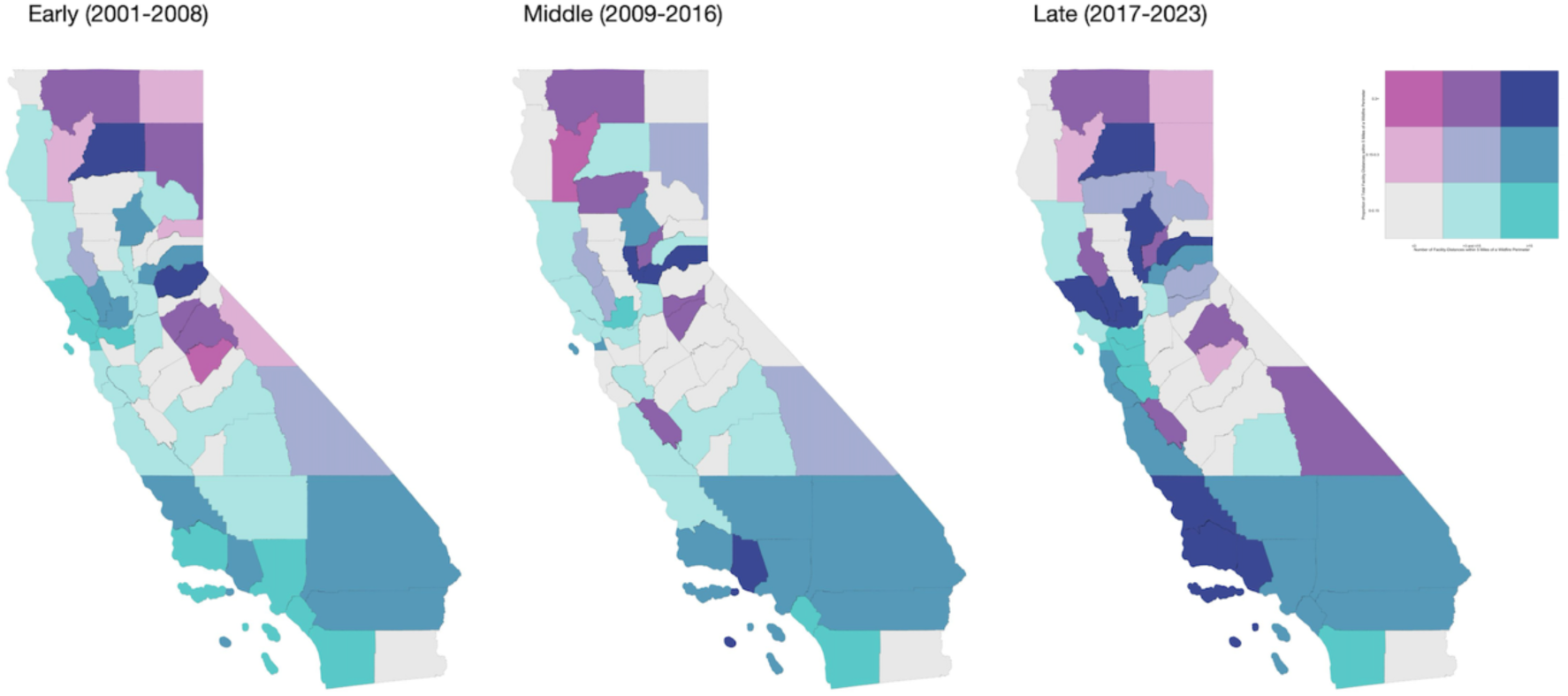
Number and Proportion of Inpatient Facilities within Five Miles of Wildfire Perimeters during the Early, Middle, and Late Time Periods. This bivariate choropleth depicts the number of facilities within each county with wildfire-facility distances ≤5 miles (aggregated into tertiles along the horizontal color palette) and the proportion of facilities within each county with wildfire-facility distances ≤5 miles (along the vertical color palette).

### Wildfire Proximity to Recently Opened Inpatient Healthcare Facilities

Over the course of the study period, 486 new inpatient facilities opened, of which 128 were hospitals and 358 were LTCFs. An analysis was performed to assess whether the opening of new healthcare facilities (which may have been sited in higher risk locations) contributed to the observed decrease in wildfire-facility distances in the late period (2017-2023) described above. Wildfire-facility distances for years 2017-2023 were separated into those representing facilities opened prior to 2017, and those opened in 2017 or later. Due to the skewed, non-normal nature of the data, wildfire-facility distances in these groups were compared via Wilcoxon rank sum test; the median wildfire-facility distance for new facilities (those first licensed and opened in 2017 or later; median 8.86 miles, IQR 7.59) was lower than for older facilities (median 12.36 miles, IQR 11.57) by 3.50 miles (W = 14312901, p < 0.0001).

### Wildfire Proximity to Healthcare Facilities Operational at the Beginning of the Study Timeframe

To assess whether the previously described decrease in wildfire-facility distances over time was due to other factors in addition to the opening of new facilities, the simple linear model described above was applied to a subset of the overall study data consisting of wildfire-facility distances for facilities that were already open at the start of the study timeframe (2001). In this model, which excluded facilities that were opened during the study period and thus excludes the effect of any facilities that were newly opened in high-risk locations, the passage of time was associated with a reduction in wildfire-facility approach distances of approximately 0.103 miles (543 feet; 166 meters) per year (p < 0.0001).

### Spatial Distribution of Wildfire Exposure at Inpatient Healthcare Facilities

The spatial distribution of facilities, wildfires, and wildfire-facility approach distances in California during the study timeframe was uneven. Inpatient healthcare facilities are concentrated in urban and peri-urban areas, particularly along the coast and in the Central Valley, with much smaller numbers of facilities located in rural and mountainous parts of the state (Supplement A).

The number and the proportion of facilities in each county experiencing wildfire approaches within 5 miles in the early, middle, and late study periods are presented in bivariate choropleths (Figure 4). In the counties of northern and northeastern California, there are comparatively few total inpatient healthcare facilities, but a high proportion of these facilities have experienced wildfire approaches within 5 miles. In contrast, a lower proportion of the inpatient healthcare facilities in counties in coastal and southern California experienced nearby wildfires, but the very large total number of facilities in these counties meant that the total number facilities threatened by wildfires within 5 miles of their location in these counties was higher than in more rural areas. The proportion of facilities and total number of facilities experiencing wildfire approaches within 5 miles was low in most counties of the Central Valley. Across California, the number of counties in which a high proportion of facilities and a high total number of facilities experienced nearby wildfires increased over the course of the study period (Figure 4, dark purple).

## DISCUSSION

Wildfire disasters in California are worsening; inpatient healthcare facility sites are increasingly exposed to this hazard. Utilizing wildfire perimeter and annual healthcare facility location data, we show that wildfires are now occurring in closer proximity to inpatient healthcare facilities, and that an increased proportion of California’s inpatient bed capacity has been exposed to nearby wildfires in recent years. Wildfire exposure distribution is uneven across California’s counties; in rural counties in northern and eastern California, a high proportion of facilities have experienced nearby wildfires, while in densely populated coastal areas, a larger total number of facilities have experienced nearby wildfires, and both absolute and proportional exposure to nearby wildfires has been low in some areas including the Central Valley.

Throughout the study period, some facilities opened, closed, or moved; the number of long-term care facilities (LTCFs) increased substantially. Analysis of recent wildfire-facility distances (2017-2023) reveals that recently licensed facilities (opened in or after 2017) were substantially more exposed than older facilities, with the median wildfire-facility distance for recently opened facilities being 3.5 miles closer than for older facilities. However, among facilities that were present in the first year of the data set (2001), decreases in wildfire facility distances were also observed over the course of the study timeframe; this increase in exposure is not attributable to the opening of new facilities during the study time frame.

Attribution of observed changes to specific causes other than the opening of new facilities was not attempted in this study, which focuses on presenting overall trends in healthcare facility hazard exposure, informing decisions about the siting of new healthcare facilities, and informing the prioritization of resources for wildfire preparedness at healthcare facilities. The observed decrease in the distances between inpatient healthcare facilities and wildfires, which remains present even when new facilities are excluded, is likely multifactorial.^2,11,17^ Development of the wildland-urban interface and impacts of climate change on wildfire risk are plausible contributors and could be topics for future analyses of the dataset assembled for this project, which is available in a public repository (https://doi.org/10.7910/DVN/MODVUK).

### Limitations

Wildfires are complex events with inherently stochastic elements. This study presents overall patterns in a large retrospective data set but cannot predict specific local risks; the absence of nearby wildfires in a given location does not mean they cannot occur. Resources including CAL-FIRE’s fire threat assessment maps are designed to provide a portrait of the spatial distribution of current wildfire risk.^13^ Some wildfires may be missing, may have been excluded, or may be inaccurately documented in the CAL-FIRE FRAP database.^15^ Changes in future fuel availability, fire weather, ignition sources, wildland management, and human settlement and facility construction patterns may be nonlinear; extrapolation of study results into the future may not have predictive value.

### Public Health and Health Policy Implications

Addressing the threat to healthcare facilities will require actions at multiple levels, including individual facilities, healthcare systems, and health policy.^18^ Goals include reducing physical risk to facilities, preparing for evacuations, and ensuring access to care.

Reducing physical risk to existing facilities involves hardening healthcare facilities and actions to reduce fire risk in the surrounding landscape. While federal guidance exists^19^ and state law requires creating defensible space around structures in fire-prone areas^20^, funding for capital investments in fireproofing hospital grounds and buildings is limited and represents a policy opportunity.

Reducing risk to new facilities requires consideration of fire threat during the siting, permitting, design, and construction process. A large fraction of the increase in wildfire exposure identified in this study is linked to newly opened inpatient healthcare facilities, predominantly long-term care facilities. Policies to ensure safe siting of future long term care facilities and design and construction practices that prioritize fire safety can help California avoid investing in new healthcare infrastructure that suffers from inherent long-term risks related due to wildfire exposure.

Wildfires create complex evacuation challenges. Acute care hospitals and LTCFs must transfer patients who may be unstable or ventilator dependent. Inpatient psychiatric facilities must maintain safety and direct observation of involuntarily hospitalized patients who may have behavioral instability or be at elevated risk of suicidal behavior. Rural facilities face lengthy transfer times that reduce evacuation efficiency.

Safe, timely evacuation of inpatient healthcare facilities requires EMS assets, availability of receiving sites, and management of resources across multiple institutions and agencies. Coordination of requests from affected facilities can help address competition for ambulances and other resources. Dedicated ambulance strike teams can augment local EMS assets.^21^ Hospital checklists can support decision-making and evacuation.^22^ Tabletop exercises, drills, investment in agencies capable of coordinating large-scale inter-facility transfers, and planning for the needs of medically vulnerable populations using resources such as HHS’s Empower dataset can also support readiness.^23^

Wildfire severity is projected to continue to rise in future years.^24^ Strategies to address the increasing risk of wildfires to healthcare infrastructure are urgently needed to protect healthcare access and ensure the well-being of individuals impacted by these events. Policies to address the root causes of the increase in wildfire exposure are also essential. It is critical that health professionals and policymakers address this imminent threat to critical healthcare infrastructure; lack of proactive efforts may lead to devastating consequences.

## CONCLUSION

California is experiencing increasingly severe wildfires and has an increasing number of inpatient healthcare facilities. Distances from inpatient healthcare facilities to nearby wildfires have decreased over the past 23 years, and an increasing proportion of inpatient beds are exposed to wildfire approaches within 5 miles of their facility. Policies to address this risk are urgently needed to protect the wellbeing of the populations served by these health systems.

## Data Availability

All data produced are available online at:
Bedi, Neil Singh; Dresser, Caleb J, 2025, "Proximity of Wildfires to Inpatient Healthcare Facilities in California, 2001-2023", https://doi.org/10.7910/DVN/MODVUK, Harvard Dataverse, V1

## SUPPLEMENTS

Higher resolution versions are available.

**Supplement A:**
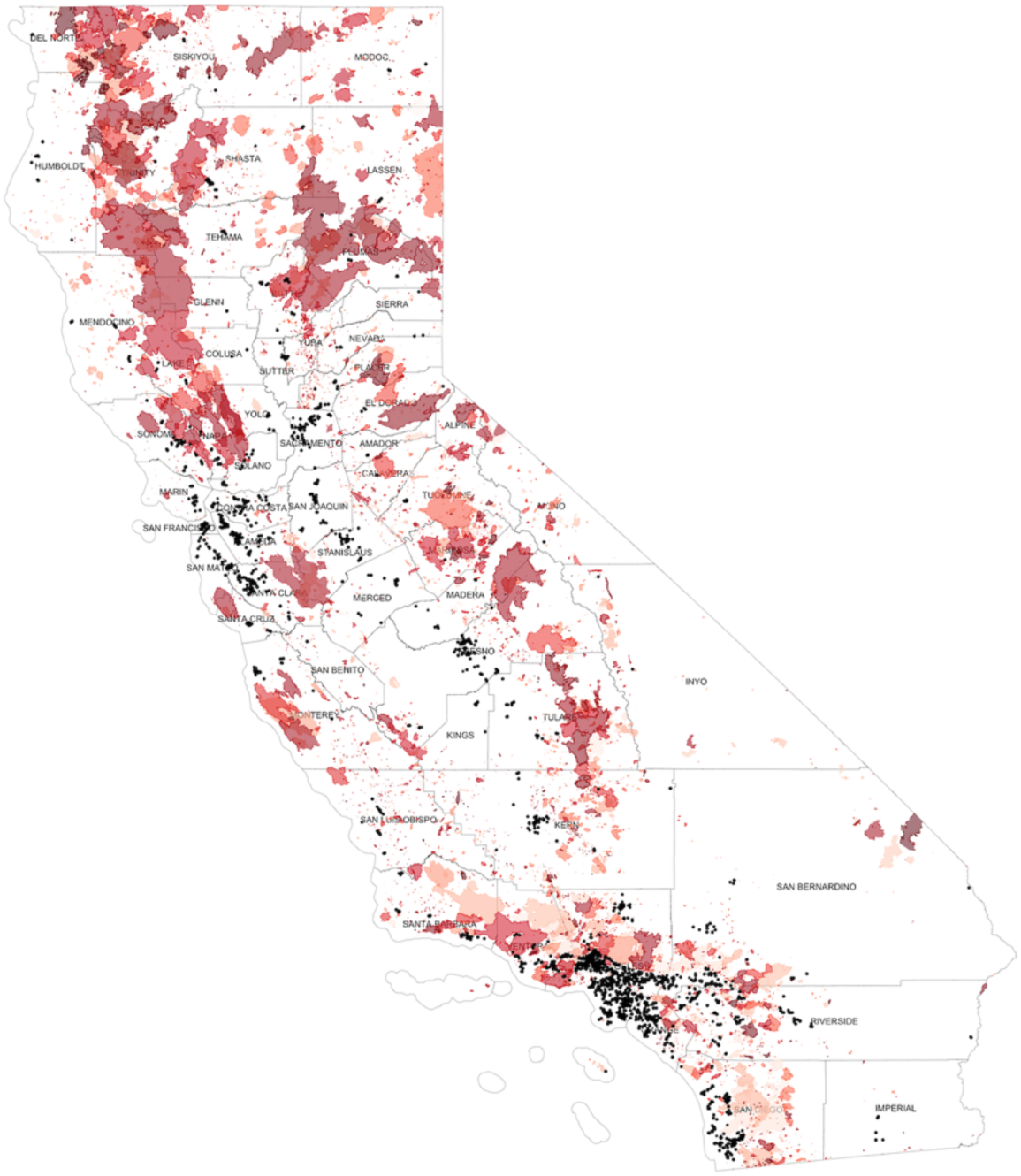
Inpatient Healthcare Facilities and Wildfires 2001 – 2023. All wildfires and inpatient facility locations used for analysis, plotted within the State of California. The polygon shading represents the year in which the fires occurred; darker shading represents more recent years. Black dots represent locations of inpatient facilities.

**Supplement B:**
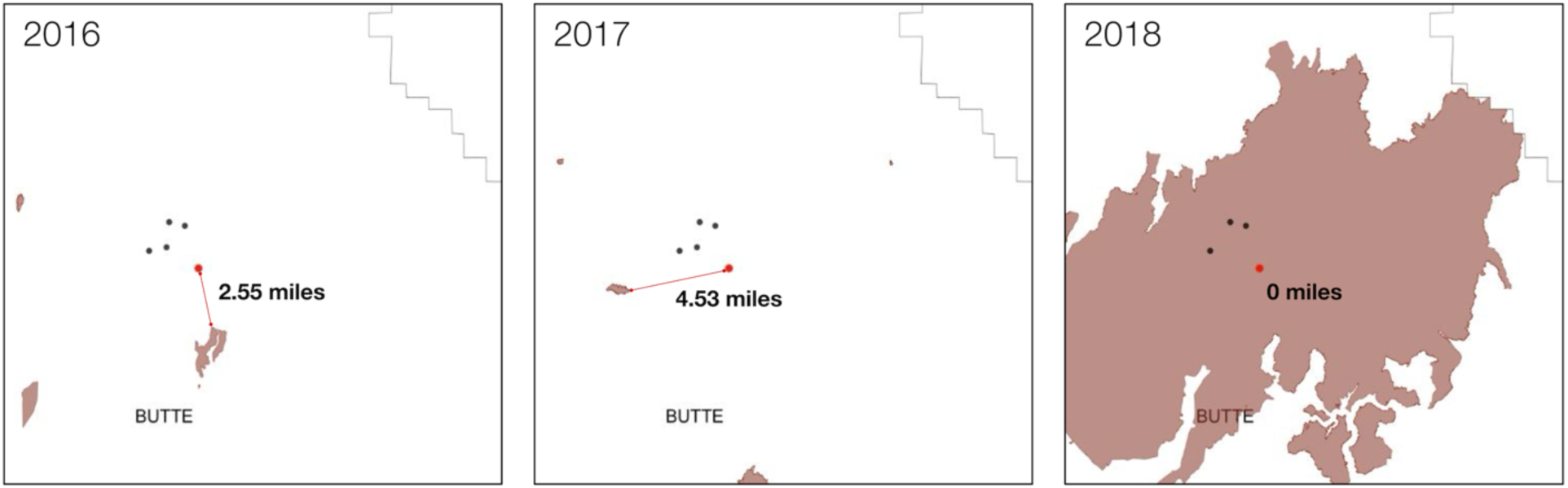
Methods Explanation: Case Series of Butte County. The panels represent three years (left, 2016; middle, 2017; right 2018) of wildfire occurrences and inpatient facility locations within Butte County. The red dot in each panel represents Adventist Health Feather River Hospital; a red line shows the distance to the nearest wildfire perimeter in each year. In 2018, the Camp Fire devastated Paradise, California, and with it, Adventist Health Feather River Hospital, which was inside the wildfire perimeter; hence, its distance for 2018 was computed as ‘0’.

**Supplement C:**
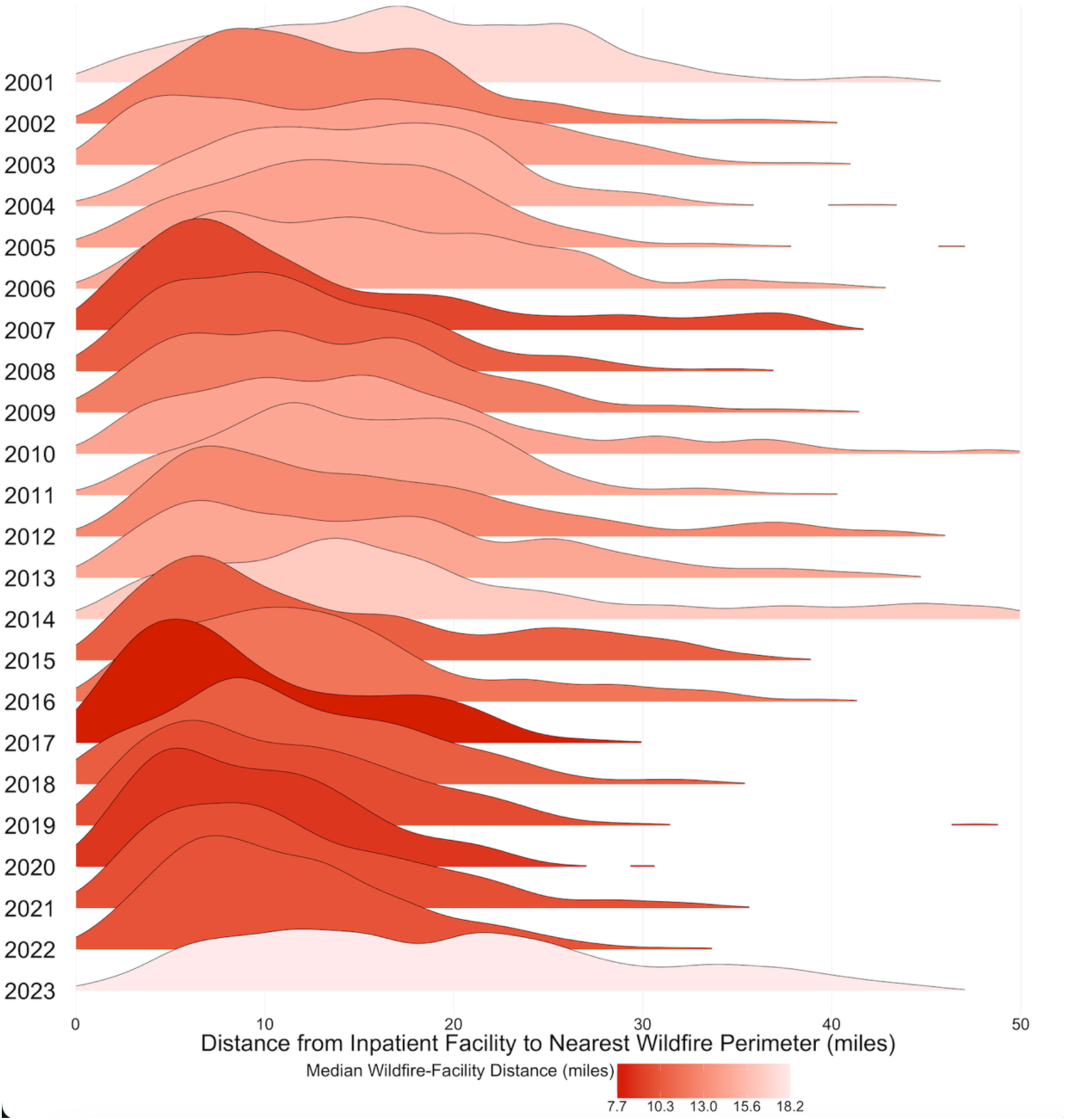
Ridgeline Plot Depicting the Annual Wildfire-Facility Distance for Inpatient Healthcare Facilities in California (2001-2023). Each distribution ridge represents the wildfire-facility distance distribution in that year. The ridges are shaded based on the median distance to the nearest wildfire perimeter; a lower median distance is indicated by a darker shade of red.

